# Familial Clustering and Genetic Analysis of Severe Thumb Carpometacarpal Joint Osteoarthritis in a Large Statewide Cohort

**DOI:** 10.1101/2022.03.03.22271851

**Authors:** Catherine M. Gavile, Nikolas H. Kazmers, Kendra A. Novak, Huong D. Meeks, Zhe Yu, Joy L. Thomas, Channing Hansen, Tyler Barker, Michael J. Jurynec

## Abstract

**Objectives:** The objectives of this study are to 1) identify individuals that required surgery for thumb carpometacarpal osteoarthritis (CMCJ OA), 2) determine if CMCJ OA clusters in multigenerational families, 3) define the magnitude of familial risk of CMCJ OA, 4) identify risk factors associated with CMCJ OA and 5) identify rare genetic variants that segregate with familial CMCJ OA.

**Methods:** We searched the Utah Population Database to identify a cohort of CMCJ OA patients that required a surgical procedure (CMC fusion or arthroplasty). Affected individuals were mapped to pedigrees to identify high-risk multigenerational families with excess clustering of CMCJ OA. Cox regression models were used to calculate familial risk of CMCJ OA in related individuals. Risk factors were evaluated using logistic regression models. Whole exome sequencing was used to identify a rare coding variant associated with familial CMCJ OA.

**Results:** We identified 550 pedigrees with excess clustering of severe CMCJ OA. The relative risk of developing CMCJ OA requiring surgical treatment was significantly elevated in first- and third-degree relatives of affected individuals, and significant associations with advanced age, female sex, obesity, and tobacco use were observed. A dominantly segregating, rare variant in *CHSY3* was associated with familial CMCJ OA.

**Conclusions:** Familial clustering of severe CMCJ OA was observed in a statewide population. Identification of a candidate gene indicates a genetic contribution to the etiology of the disease. Our data indicate the genetic and environmental factors contribute to the disease process, further highlighting the multifactorial nature of the disease.

**Key messages:**

- We study a unique cohort of individuals requiring surgical management of CMCJ OA.
- Severe CMCJ OA clusters in large, multigenerational families indicating a genetic contribution to the disease.
- .We discovered a dominant coding variant in CHSY3 in a family with severe CMCJ OA.

## Introduction

Hand osteoarthritis (OA) is the most common form of OA (1, 2). It is a debilitating disease that comprises a significant healthcare burden. The different subsets of hand OA are categorized based on the joints within the hand that are affected. The thumb carpometacarpal joint (CMCJ) is the saddle joint between the trapezium and the first metacarpal, which allows for a wide range of motion and is critical for normal hand function, albeit with the caveat that it is also easily damaged (3, 4). Although CMCJ OA is common, little is known about the biological mechanisms that are responsible for disease onset and progression. Currently, treatment of this disease is limited to surgery and measures that temporarily alleviate pain. The risk factors for developing CMCJ OA in cohorts defined by diagnostic codes or radiographic evidence include age (3), sex (5-7), and hypermobility (8, 9). Additionally, a recent study found that being overweight (men) and obesity (both sexes) are risk factors associated with CMCJ OA despite the joint not being a weight-bearing unit (10).

Radiographic evaluation is the primary means of diagnosing CMCJ OA. However, radiographic severity does not correlate with clinical severity(5), and not all patients with a radiographic diagnosis have a clinical phenotype with pain or hand dysfunction (11). Therefore having the diagnosis does not necessarily lead to the development of symptoms that require surgical treatment or surgical intervention. Identification of individuals requiring surgical treatment for CMCJ OA captures those with potentially severe symptoms/disease, and allows higher resolution to identify risk factors and genes that contribute to pathologic forms of CMCJ OA.

There are at present no drugs that specifically target the progression of CMCJ OA since few of the pathways and genes that drive the disease have been identified (12, 13). Several studies have indicated that CMCJ OA is heritable in closely related individuals (14-16) and others have identified several risk loci and candidate genes (17-22). Given that few genes have been associated with CMCJ OA, there is clearly a need to identify more pathways that have a strong contribution to disease susceptibility. One way to identify genes with a determinant effect on disease is to study families that have severe forms of disease (23-27). Our criteria to identify a unique CMCJ OA cohort with potentially severe disease (requiring surgical management and excluding confounding factors) makes identifying large, multigenerational families unambiguous.

Here we employed a unique resource, the Utah Population Database (UPDB), to 1) identify individuals with severe CMCJ OA (requiring surgery), 2) determine if severe CMCJ OA clusters in large, multigenerational families, 3) define the magnitude of familial risk of CMCJ OA, 4) identify risk factors associated with CMCJ OA and quantify the magnitude of risk, and 5) discover rare genetic variants that segregate with severe familial CMCJ OA.

## Methods

### Study approval

The Institutional Review Boards of the University of Utah (IRB # 79442) and Intermountain Healthcare (IRB # 1050554), and the Resource for Genetic and Epidemiologic Research approved this study.

### The Utah Population Database (UPDB)

Our study utilizes data drawn from UPDB (https://uofuhealth.utah.edu/huntsman/utah-population-database/). The UPDB contains data on over 11 million individuals from the late 18th century to the present. Data are updated as they become available from statewide birth and death certificates, hospitalizations, ambulatory surgeries, and drivers’ licenses. UPDB creates and maintains links between the database and the medical records held by the two largest healthcare providers in Utah as well as Medicare claims.

### Selection of cases and controls

We identified individuals with severe CMCJ OA between 1996 and 2020 in the UPDB using the following diagnosis and related procedure codes: diagnostic codes for CMCJ OA - ICD-9 715·04 or 715·14; ICD-10 M18·0 or M18·1x or M19·04; procedure codes for CMC arthroplasty and fusion - ICD-9 8269, 8174, or 8175; CPT 25447 or 2544. Inclusion as a case required presence of one diagnostic code and one procedure code. Individuals were excluded if they had a traumatic event to the thumb CMC joint, inflammatory arthritis, and ligamentous hyperlaxity. Controls were age and sex matched and were selected if they had no history of a CMCJ OA diagnosis or related procedure, and excluded based on criteria mentioned above (Supplemental Data). Affected individuals were required to have relatives in the UPDB to be included in our study cohort so we could link them to pedigrees. Manual chart review was performed on a random set of patients to verify our coding strategy to identify CMCJ OA cases (Supplemental Data).

### High-risk pedigree identification

To determine if there was excess familial clustering of severe CMCJ OA in each pedigree, we utilized a threshold of ≥ 2·0 for the Familial Standardized Incidence Ratio (FSIR). FSIR allows for the quantification of familial risk of a disease by comparing the incidence of a disease in a family to its expected incidence in the general population (Supplemental Data) (25, 28).

### Familial risk analysis

Baseline demographic characteristics of the relatives of CMCJ OA patients and their matching controls were compared using t-tests for continuous variables and chi-square tests for categorical variables. Familial risk of CMCJ OA was estimated using Cox regression standard competing risk models to assess risk in first-degree relatives (FDRs), second-degree relatives (SDRs), and third-degree relatives (TDRs) of CMCJ OA patients. Specific relatives (FDRs, SDRs, and TDRs) of CMCJ OA patients were compared with the relatives of their matching controls. Separate hazard ratios were estimated for each relationship type overall and by sex. Covariates in the Cox regression models include sex, birth year, Caucasians, and Hispanic. Time was measured in years, and individuals were first followed from birth or 01/01/1996, whichever occurred later, and right-censored at the time of death, or CMCJ OA diagnosis, or 12/31/2020, whichever occurred first. All known relatives of patients and their matching controls were included in the analyses, even if that relative had been previously counted (28). Huber-White sandwich estimator of variance of regression parameters in the Cox models were used to correct for the non-independence of observations within families (29). Family members of controls were used as the reference group in all analysis. All statistical tests were two-sided, and a P-value <0·05 was considered statistically significant.

### Risk factor analysis

Baseline demographic characteristics of CMCJ OA patients and their matching controls were compared using t-tests for continuous variables and chi-square tests for categorical variables. Risk factors (Supplemental Table S1 for ICD-9/10 codes) were assessed using conditional logistic regression models. All statistical tests were two-sided, and a P-value <0·05 was considered statistically significant. Odds ratios and 95% CI were calculated.

### Age- and sex-specific standardized incidence rates

All individuals included in the study had a known birth year, informative sex, and resided in Utah any time between 1996 and 2020. Demographic characteristics of CMCJ OA patients and the unaffected population were compared using t-tests for continuous variables and chi-square tests for categorical variables. Age-standardized incidence rates by sex were calculated using the direct method. Person-years were calculated for CMCJ OA patients and the unaffected population. Each patient contributed 1 person-year for every year lived in Utah from 1996 until first diagnosis with CMCJ OA. Each unaffected individual contributed 1 person year for every year lived in Utah from 1996 to 2021. The female-to-male incidence ratios and 95% confidence intervals were estimated assuming log-normal distribution for 9 age groups.

### Whole exome sequencing and analysis

Whole exome sequencing (WES) and analysis was performed as previously described (Supplemental Data) (24).

## Results

### Identification and demographic detail of the severe CMCJ OA cohort

We applied a strict definition of CMCJ OA to identify individuals with potentially severe forms of CMCJ OA. We define ‘severe’ CMCJ OA as having been diagnosed and undergone a surgical procedure (CMC fusion or arthroplasty). Using the described coding strategy (Methods and Supplemental Data), we searched 7,311,712 UPDB records and identified 4707 individuals with severe CMCJ OA. Mean birth year of the CMCJ OA cohort was 1950 (± 10·3), 76·1% were female, and 86·9% of individuals were white (Supplemental Table S2). We performed manual chart review on a random subset of 25 individuals to verify the CMCJ OA diagnosis (see Supplemental Data and Supplemental Figure S1), and found that all 25 individuals had a verifiable CMCJ OA diagnosis confirmed by radiographic evidence.

### Identification of high-risk pedigrees

To test if there is significant familial clustering of severe CMCJ OA in our cohort, we analyzed cases with severe CMCJ OA that linked to a pedigree using the Familial Standardized Incidence Ratio (FSIR) calculation(28). We identified 550 unrelated, multigenerational, high-risk pedigrees that had at least 4 living members in the UPDB and an increased clustering of severe CMCJ OA, defined by a FSIR ≥ 2.0 (p-value < 0·05). Of the 550 high-risk pedigrees, the FSIR ranged from 2·03 – 21·08 (mean 3·4 ± SD 1·8). Founder birth year, number of descendants, number of affected individuals, and FSIR values are indicated for 10 representative high-risk pedigrees in Table 1. Figure 1 illustrates two multigenerational high-risk pedigrees. One pedigree has at least 20 known affected individuals and a FSIR of 2·1 (Figure 1A) and another has at least 4 known affected individuals and a FSIR of 4·6 (Figure 1B). The identification of high-risk pedigrees indicates significant familial clustering of severe CMCJ OA in our cohort.

**Table 1.** High-Risk Pedigrees with Excess Familial Clustering of Thumb Carpometacarpal Joint Osteoarthritis. The familial standardized incidence ratio (FSIR) and p-values were calculated according to the method of Kerber(28).

| Founder Birth Year | Number of<br>Descendants | Number of<br>Affected<br>Individuals | FSIR<sup>#</sup> |
| --- | --- | --- | --- |
| 1766* | 22,460 | 20 | 2.1 |
| 1740 | 22,450 | 24 | 2.1 |
| 1788 | 16,344 | 19 | 2. |
| 1761 | 11,909 | 15 | 3.7 |
| 1754 | 5,105 | 12 | 4.1 |
| 1788 | 4,628 | 8 | 6.3 |
| 1815 | 1,127 | 7 | 7.8 |
| 1783 | 2,228 | 6 | 9.1 |
| 1809 | 979 | 5 | 12.5 |
| 1848 | 525 | 4 | 21.1 |

\*indicates pedigree represented in Figure 1A. # indicates that all FSIR P-values are $P < 0.05$ .

**Figure 1.**
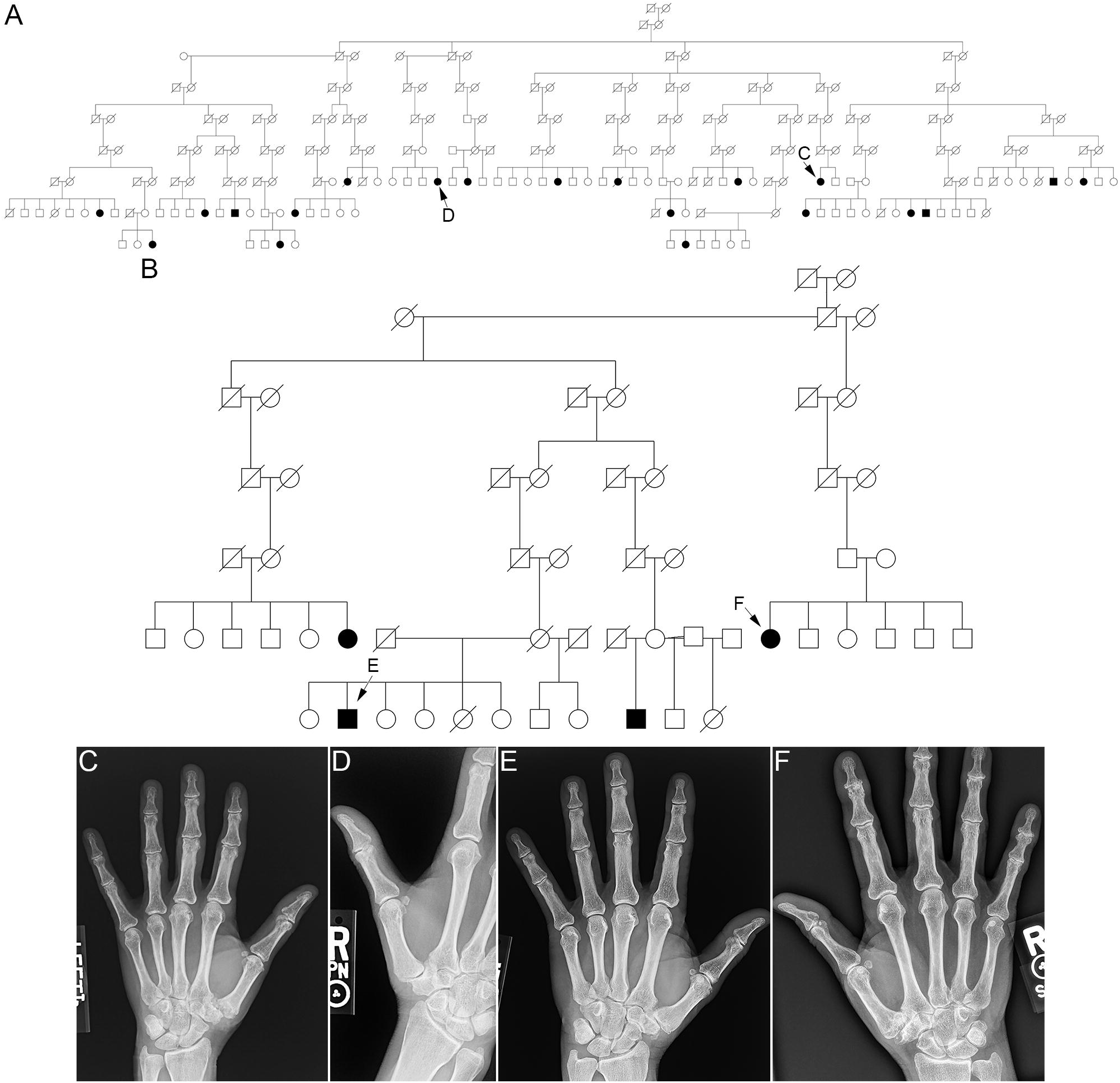
High-risk thumb carpometacarpal joint osteoarthritis pedigrees. (A,B) Examples of two high-risk pedigrees segregating severe CMCJ OA identified from the Utah Population Database. Circles = females, squares = males, slash = deceased. Filled circles/squares = affected individuals; open circles/squares = individuals with unknown affection status. C, D E, and F indicate subjects for whom radiographs are shown. FSIR of pedigree A is 2.1 and pedigree B is 4.6. (C-F) Hand radiographs of individuals in the pedigrees shown in A and B.

### Familial risk

To determine whether there is an increased risk of CMCJ OA among closely related individuals, we examined the relative risk of developing CMCJ OA in first-, second-, and third-degree relatives (demographics of the population listed in Supplemental Table S3). The risk of developing CMCJ OA was approximately 3·66-fold greater in first-degree relatives of both sexes of patients compared to controls (RR, 3·66 [95% CI, 2·73 - 4·9], P < 0.001), while males and females had a different magnitude of risk; males: RR= 6·26 [95% CI 3·72-10·56], P < 0.001; females: RR=3·17 [95% CI 2.30-4·36], P < 0.001) (Table 2). We were unable to detect a significant elevated risk of CMCJ OA in second-degree relatives. For third-degree relatives, we saw a slight increase in relative risk for CMCJ OA development in both sexes (RR 1·19 [95% CI, 1·01-1·40], P = 0·043), but more significantly in males (RR=1·39 [CI 1·02-1·90], P = 0·038) (Table 2). Together with the familial clustering of severe CMCJ OA, these data indicate a genetic component to the disease.

**Table 2.** Increased Familial Risk of Thumb Carpometacarpal Joint Osteoarthritis.

**Both Sexes**
|  | N of relatives of CMCJ OA cases | N of relatives of matching controls | P-value | Relative Risk | 95% CI Lower Limit | 95% CI Upper Limit |
| --- | --- | --- | --- | --- | --- | --- |
| Had a FDR with CMC | 145 | 195 | 0.000 | 3.66 | 2.73 | 4.90 |
| Had a SDR with CMC | 44 | 167 | 0.197 | 1.25 | 0.89 | 1.76 |
| Had a TDR with CMC | 207 | 775 | 0.043 | 1.19 | 1.01 | 1.40 |

**Male**
|  | N of relatives of CMCJ OA cases | N of relatives of matching controls | P-value | Relative Risk | 95% CI Lower Limit | 95% CI Upper Limit |
| --- | --- | --- | --- | --- | --- | --- |
| Had a FDR with CMC | 39 | 30 | 0.000 | 6.26 | 3.72 | 10.56 |
| Had a SDR with CMC | 7 | 30 | 0.808 | 1.11 | 0.48 | 2.55 |
| Had a TDR with CMC | 60 | 190 | 0.038 | 1.39 | 1.02 | 1.90 |

**Female**
|  | N of relatives of CMCJ OA cases | N of relatives of matching controls | P-value | Relative Risk | 95% CI Lower Limit | 95% CI Upper Limit |
| --- | --- | --- | --- | --- | --- | --- |
| Had a FDR with CMC | 106 | 165 | 0.000 | 3.17 | 2.30 | 4.36 |
| Had a SDR with CMC | 37 | 137 | 0.192 | 1.28 | 0.88 | 1.86 |
| Had a TDR with CMC | 147 | 585 | 0.256 | 1.12 | 0.92 | 1.35 |

FDR = first-degree relatives; SDR = second-degree relatives; TDR = third-degree relatives

### Age-standardized sex-specific incidence rates of severe CMCJ OA

OA in general is more prevalent in females, and the same has been reported for CMCJ OA, with almost three quarters of affected individuals identified as female (1, 2, 30). To determine if there is an age and sex bias associated with severe CMCJ OA, we examined age-standardized sex-specific incidence rates in our cohort from 1996-2020. We found a significant association of sex and age with severe CMCJ OA in our cohort. Of 6,083,729 unaffected individuals and 4,540 CMCJ OA cases, a higher proportion of cases were female (76·3% of CMCJ OA cases versus 48·7% of controls), and were older (mean ± SD birth year 1950·3 ± 10·3 for CMCJ OA cases and 1980·2 ± 24·6 for controls) (P <0·001) (Supplemental Table 4). We also determined that females had a significantly higher rate of severe CMCJ OA from the ages of 30–89 years compared to males, with the highest female-to-male incidence ratio being 4·68 (95% CI 4·07–5·37) in the age group 50–59 years (Table 3). Our results indicate that being female of advanced age is a significant risk factor for severe CMCJ OA.

**Table 3.**
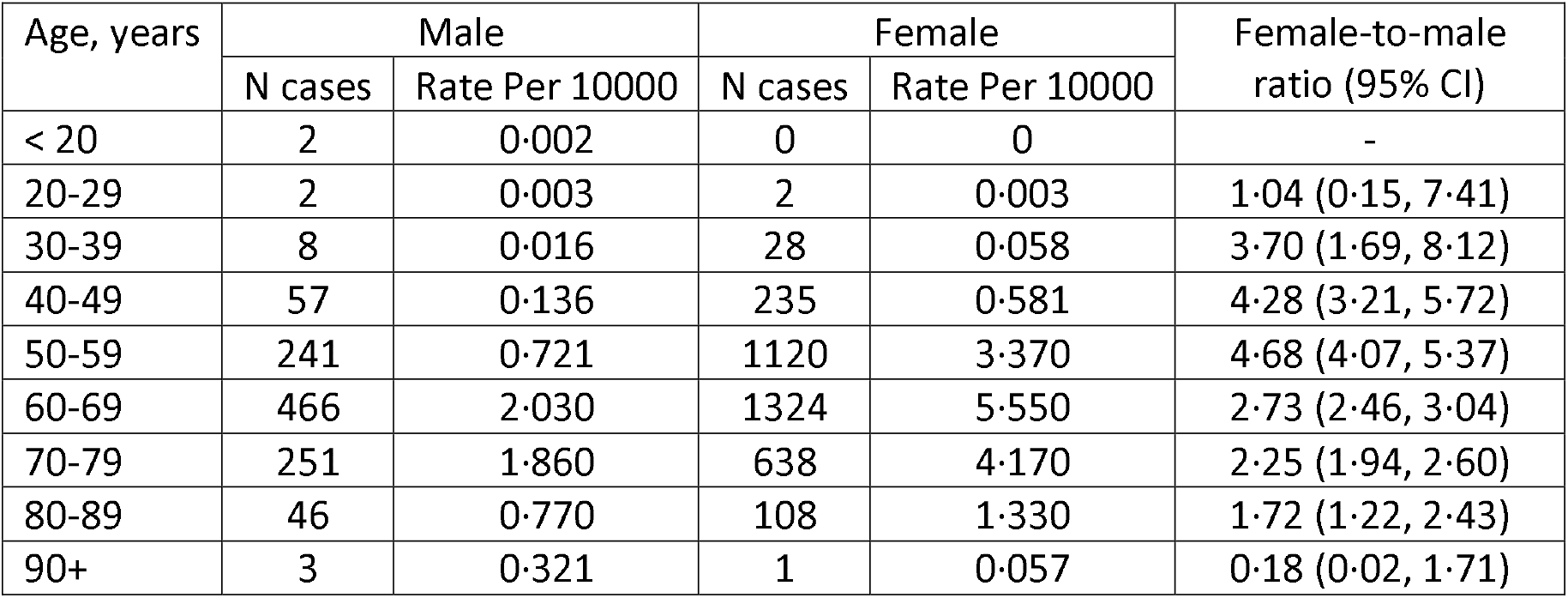
Age-Specific Incidence Rates of Thumb Carpometacarpal Joint Osteoarthritis by Sex and Female-to-Male Incidence Ratios.

### Risk factors associated with severe CMCJ OA

The risk factors that contribute to severe CMCJ OA are unknown. We analyzed several risk factors to determine if they are associated with severe CMCJ OA in our cohort. We focused on risk factors that have previously been linked to osteoarthritis and hand OA (see Supplemental Table S1 for risk factor diagnostic codes) (30-32). We examined the association of severe CMCJ OA with obesity, diabetes, tobacco use, alcohol use, and being related to an individual diagnosed with and surgically treated for CMCJ OA (to the third degree) in the same cohort used for the age- and sex-specific analysis. We examined the relative risk of the above risk factors independently in males and females while adjusting for demographic features (race and ethnicity), and diabetes (see Methods and Supplemental Table 3). When adjusting for diabetes, obesity was identified as a risk factor for males (RR of 1·59 [95% CI 1·36-1·86]) and females (RR 1·27 [95% CI 1·16-1·38])· Tobacco use was a risk factor for both sexes (males: RR 1·29 [95% CI 1·12-1·49]; females: RR 1·45 [95% CI 1·33-1·59]) (Table 4). Independent of diabetes, having a first-degree relative (males: RR 3·73 [95% CI 2·39-5·83]; females: RR 3·26 [95% CI 2·50-4·26]) or a third-degree relative (males: RR 1·62 [95% CI 1·18-2·23]) with severe CMCJ OA was a significant risk factor (Table 4). We find the same risk factors when not adjusting for diabetes (Table 4). Our analyses indicate that Hispanics are slightly less likely to develop CMCJ OA (RR 0·85 [95% CI 0·78-0·93] - non-white vs white for both sexes, irrespective of adjustment for diabetes) (Supplemental Table S5). These data indicate that obesity, diabetes, tobacco use, and having a first- and third-degree relative with severe CMCJ OA are all significant risk factors for severe CMCJ OA.

**Table 4.** Risk Factors Associated with of Thumb Carpometacarpal Joint Osteoarthritis.

**Males – adjusted for diabetes**
|  | CMCJ<br>OA<br>cases | Controls | P-value | Relative<br>Risk | 95% CI<br>Lower Limit | 95% CI<br>Upper Limit |
| --- | --- | --- | --- | --- | --- | --- |
| Ever diagnosed with alcoholism | 55 | 340 | 0.110 | 0.78 | 0.57 | 1.06 |
| Ever diagnosed with diabetes | 348 | 1571 | 0.414 | 1.07 | 0.91 | 1.24 |
| Ever diagnosed with obesity | 327 | 1169 | 0.000 | 1.59 | 1.36 | 1.86 |
| Ever diagnosed with tobacco | 417 | 1862 | 0.000 | 1.29 | 1.12 | 1.49 |
| Had at least one FDR with CMCJ<br>OA | 36 | 49 | 0.000 | 3.73 | 2.39 | 5.83 |
| Had at least one TDR with CMCJ<br>OA | 55 | 180 | 0.003 | 1.62 | 1.18 | 2.23 |

**Females – adjusted for diabetes**
|  | CMCJ<br>OA<br>cases | Controls | P-value | Relative<br>Risk | 95% CI<br>Lower Limit | 95% CI<br>Upper Limit |
| --- | --- | --- | --- | --- | --- | --- |
| Ever diagnosed with<br>alcoholism | 113 | 471 | 0.293 | 1.12 | 0.90 | 1.40 |
| Ever diagnosed with diabetes | 843 | 4469 | 0.045 | 0.91 | 0.83 | 1.00 |
| Ever diagnosed with obesity | 1104 | 4900 | 0.000 | 1.27 | 1.16 | 1.38 |
| Ever diagnosed with tobacco | 952 | 3778 | 0.000 | 1.45 | 1.33 | 1.59 |
| Had at least one FDR with<br>CMCJ OA | 96 | 151 | 0.000 | 3.26 | 2.50 | 4.26 |
| Had at least one TDR with<br>CMCJ OA | 132 | 616 | 0.554 | 1.06 | 0.87 | 1.29 |

**Males only – not adjusted for diabetes**
|  | CMCJ<br>OA<br>cases | Controls | P-value | Relative<br>Risk | 95% CI<br>Lower Limit | 95% CI<br>Upper Limit |
| --- | --- | --- | --- | --- | --- | --- |
| Ever diagnosed with<br>alcoholism | 55 | 340 | 0.109 | 0.78 | 0.57 | 1.06 |
| Ever diagnosed with obesity | 327 | 1169 | 0.000 | 1.62 | 1.40 | 1.89 |
| Ever diagnosed with tobacco | 417 | 1862 | 0.000 | 1.30 | 1.12 | 1.49 |
| Had at least one FDR with<br>CMCJ OA | 36 | 49 | 0.000 | 3.73 | 2.39 | 5.83 |
| Had at least one TDR with<br>CMCJ OA | 55 | 180 | 0.003 | 1.62 | 1.18 | 2.23 |

**Females – not adjusted for diabetes**
|  | CMCJ<br>OA<br>cases | Controls | P-value | Relative<br>Risk | 95% CI<br>Lower Limit | 95% CI<br>Upper Limit |
| --- | --- | --- | --- | --- | --- | --- |
| Ever diagnosed with<br>alcoholism | 113 | 471 | 0.281 | 1.13 | 0.91 | 1.40 |
| Ever diagnosed with obesity | 1104 | 4900 | 0.000 | 1.23 | 1.14 | 1.34 |
| Ever diagnosed with tobacco | 952 | 3778 | 0.000 | 1.45 | 1.33 | 1.58 |
| Had at least one FDR with<br>CMCJ OA | 96 | 151 | 0.000 | 3.26 | 2.50 | 4.26 |
| Had at least one TDR with<br>CMCJ OA | 132 | 616 | 0.548 | 1.06 | 0.87 | 1.29 |

FDR = first-degree relatives; SDR = second-degree relatives; TDR = third-degree relatives. All conditions are yes vs. no.

### Chondroitin Synthase-3 is a candidate CMCJ OA gene

A major limitation to the development of disease modifying OA therapies is a lack of targets. To identify genes that are major risk factors for development of severe CMCJ OA, we analyzed the exomes of one unaffected and five affected individuals from a family in which CMCJ OA segregated as an apparent autosomal dominant trait (Figure 2a, b). Following our previously published genomic analysis pipeline (24) using pVAAST in conjunction with Phevor2 and ANNOVAR, we identified a rare coding variant in Chondroitin Synthase-3 (PVAAST: p-value = 4·7×10^−5^; LOD 1·5; PHEVOR score: 4·1, final rank = 5) (*CHSY3*; NM_175856: exon3:c.G1885A:p.G629R, rs145272862, MAF 0·0001312) (Figure 2c). The rs145272862 SNP results in the non-synonymous substitution of an invariant glycine (in the vertebrate lineage) to an arginine in the chondroitin N-acetylgalactosaminyltransferase domain of CHSY3. CHSY3 is a glycosyltransferases involved in initiation and elongation of the of the chondroitin sulfate glycosaminoglycan side chains on a core protein (33), including the major chondroitin sulfate proteoglycan of cartilage, Aggrecan (ACAN). Identification of a CHSY3 coding variant in a CMCJ OA family suggests that alterations in chondroitin sulfate proteoglycans, which include ACAN, may be a major risk factor for CMCJ OA.

**Figure 2.**
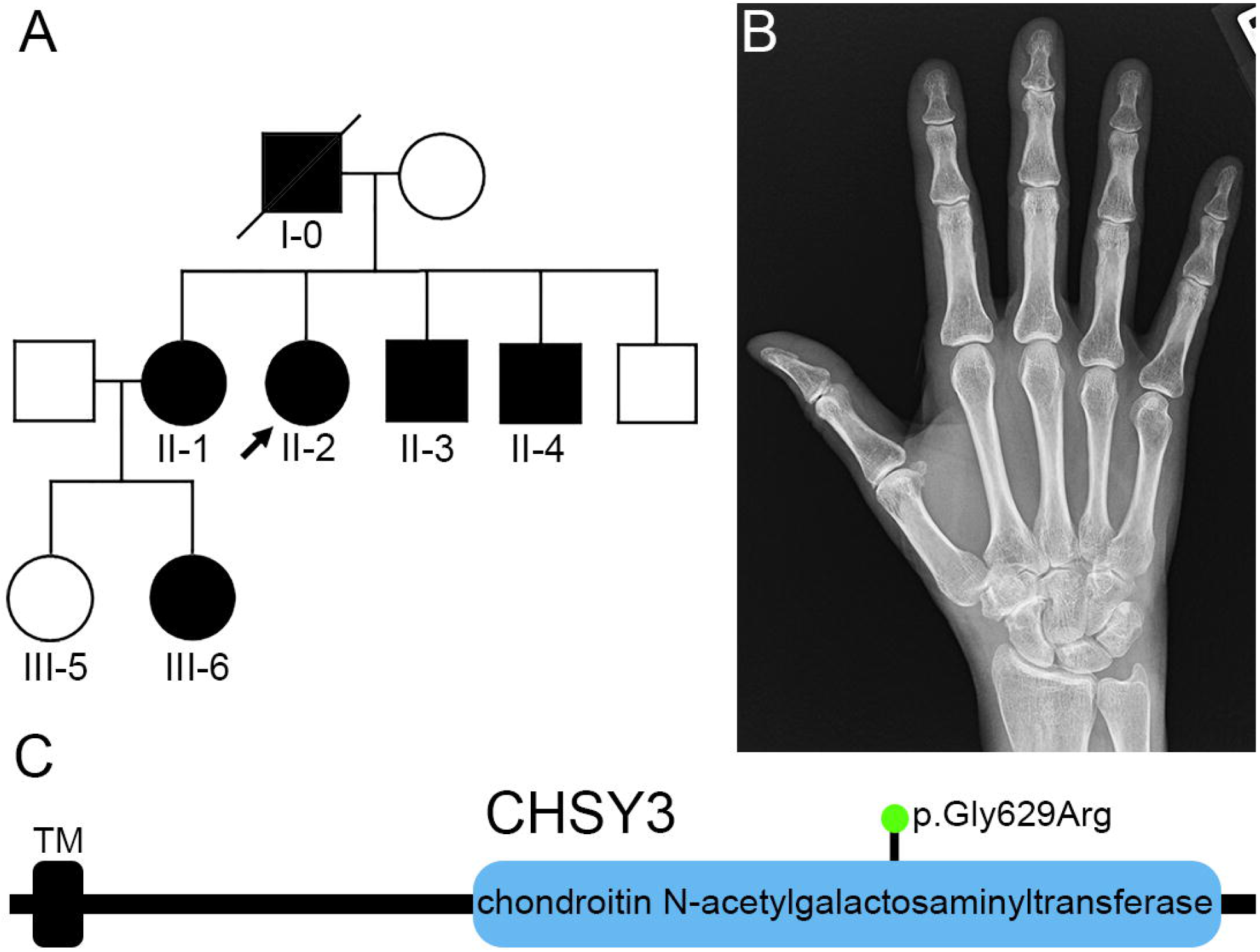
A dominant *CHSY3* mutation segregates with thumb carpometacarpal joint osteoarthritis. (A) Family 1 pedigree. Severe CMCJ OA segregates as an apparent autosomal dominant trait. Arrow marks the proband (II-2). The founder (deceased, I-0) was not genotyped, but had a medical history of CMCJ OA. Exomes were sequenced from 2 generations of the family (individuals II-1-4 and III-5 and 6). All affected individuals genotyped (II-1-4 and III-6) were heterozygous for the rare variant (rs145272862) and the unaffected individual (III-5) genotyped was homozygous for the reference allele. (B) Right hand radiograph of the proband demonstrating CMCJ OA. (C) Schematic diagram of the CHSY3 protein indicating the transmembrane domain (TM) and the chondroitin N-acetylgalactosaminyltransferase domain and the location of the p.Gly629Arg mutation in the catalytic domain.

## Discussion

In this study, we leveraged a unique statewide medical genetics resource, the UPDB, to identify a cohort of individuals diagnosed with severe CMCJ OA. Previous studies primarily focused on CMCJ OA cohorts defined by radiographic evidence of OA, which does not correlate with symptomatic severity (5, 11). Our study focuses on a unique population of CMCJ OA individuals; those with symptoms severe enough to require surgical management of symptoms (CMC fusion or arthroplasty). From this cohort of individuals with severe CMCJ OA we have i) identified 550 unrelated high-risk pedigrees demonstrating familial enrichment severe CMCJ OA, ii) determined that first- and third-degree relatives of an individual with severe CMCJ OA is at approximately 3·66-fold and 1·19-fold increased risk of developing the disease, respectively, iii) determined that age, sex, obesity, tobacco-use, and having a first- or third-degree relative with severe CMCJ OA are significant risk factors associated with the disease, and iv) and identified a rare, dominantly segregating coding variant in *CHSY3* as a candidate gene for CMCJ OA. In sum, these data suggest that both genetic and physiological factors contribute to the development of severe CMCJ OA in a large population-based cohort.

The risk factors in our surgical CMCJ OA cohort is consistent with other studies that have examined risk factors for CMCJ OA in other populations (2, 5-7, 9, 10, 16). Although prior studies have associated tobacco use with less hand OA (31), to our knowledge this is the first-time tobacco use has been associated with increased risk of CMCJ OA. It is unclear what the mechanistic link is between tobacco use and surgery in our cohort. Awareness of these comorbidities may help guide the clinical diagnosis of this condition in at-risk populations, particularly those with affected family members, and assist in the identification of individuals who are unlikely to do well with non-operative treatment. The mechanisms by which environmental and physiological risk factors interact with an individual’s genetic background to contribute to CMCJ OA remains to be elucidated.

CMCJ OA is highly heritable (14-16), yet few coding variants have been definitively linked to CMCJ OA (17, 18, 20-22, 34). Using the UPDB enabled us to discover a rare coding variant in a high-risk pedigree, a potentially powerful way to define pathways with a determinant effect on disease development (23, 24, 35, 36). To our knowledge, our study is the first to identify a large number of multigenerational severe CMCJ OA pedigrees, determine relative risk among relatives and identify a coding variant associated with CMCJ OA. Identification of causal gene variants in these families will inform us about pathways that contribute to CMCJ OA when disrupted.

To begin to define these pathways, we performed genomic analyses on a high-risk pedigree and identified a rare, dominantly segregating coding variant (rs145272862, c.G1885A:p.G629R) in *CHSY3*. CHYS3 is a glycosyltransferases that initiates and adds chondroitin sulfate glycosaminoglycan side chains to a core protein to from a functional proteoglycan (33). The variant is predicted to be damaging and is located in a highly conserved amino acid in the enzymatic domain of CHSY3 (Figure 2), although functional studies are needed to define the effect of the variant on protein function. One hypothesis is the disease allele reduces chondroitin N-acetylgalactosaminyltransferase activity of CHSY3, thereby limiting initiation or elongation of chondroitin sulfate glycosaminoglycan side chains to the core protein. Aggrecan (ACAN) is the major chondroitin sulfate proteoglycan in cartilage, and loss of chondroitin sulfate concentration and reduction of side chain length is associated with OA severity (37). A reduction in CHSY3 activity may alter the chondroitin sulfate glycosaminoglycan content of ACAN, thereby leading to the initiation of cartilage catabolism and onset of OA. Consistent with this, mice lacking Chsy3 display intervertebral disc degeneration, including loss of *Acan* expression and increased expression of catabolic factors in nucleus pulposus tissue (38). Modification of the chondroitin core protein is a recurrent risk factor for OA as loci containing other enzymes (CHST11 and CHST2) in the pathway have been identified in OA GWAS (20, 39), making this pathway an appealing candidate for therapeutic intervention. Genomic analysis of additional independent families will determine if *CHSY3* (or other pathway members) is recurrently disrupted in CMCJ OA. Characterization of the disease allele in animal models will provide insight into the biological mechanism of how disruption of CHSY3 activity leads to the onset and progression of OA.

Our study has several limitations. Our definition of ‘severe’ CMCJ OA (requiring surgical intervention) may be dependent on several factors. There are no concrete indications for CMCJ OA surgery other than pain and disability that is severe enough (in the patient’s perspective), alongside ‘failure’ of conventional nonoperative treatment (splinting > 6 weeks, NSAIDs, steroid injection). In addition, there is conflicting literature regarding the relationship between radiographic severity and level of pain and disability. While it is possible that disease/radiographic severity and clinical severity may not be related, this definition allows us to focus on a specific CMCJ OA cohort that has not previously been studied and makes identifying families with the phenotype unambiguous. The relative risk and FSIR calculations are likely underestimates for CMCJ OA due to several factors. First, our cohort was limited to individuals identified through medical coding and our analysis can only identify individuals diagnosed in Utah. We are thus missing individuals diagnosed out of state and affected individuals who have not sought medical care. Because of these factors, our analyses are likely an underrepresentation of CMCJ OA, and in high-risk pedigrees we consider individuals without a CMCJ OA diagnosis as ‘affection status unknown’ until we can definitively determine if they are unaffected or affected. We have a small number of ethnic minorities in our study, which reflects the overall population of Utah. While we detected significant differences in risk factors in this population, given the small number of individuals we are cautious not to over interpret this data until a larger cohort can be studied.

In sum, our data indicate that both genetic and physiological factors contribute to the development of severe CMCJ OA in a large population-based cohort. We can use high-risk pedigrees to identify genes and pathways that are major susceptibility factors, which may provide significant insight into therapeutic intervention and may be useful biomarkers for early diagnosis for CMCJ OA.

## Data Availability

All data produced in the present study are available upon reasonable request to the authors

## Contributors

CMG, NHK, and MJJ conceived and designed the study, analyzed data, and wrote the manuscript. HM and KAN analyzed data and contributed to writing of the manuscript. ZY, JLT, CH, and TB analyzed data and provided feedback on the manuscript.

## Data Sharing

Data are available upon request.

## Acknowledgements

We would like to thank the individuals who participated in this research.

## Figure Legends

**Supplemental Figure S1** – Hand Radiographs of Individuals in the Thumb Carpometacarpal Joint Osteoarthritis Cohort. Each image represents a unique individual.

**Supplemental Table S1** – Identification of Risk Factors Using Diagnostic Coding.

**Supplemental Table S2** – Demographic Characteristics of Individuals Diagnosed with Thumb Carpometacarpal Joint Osteoarthritis and Matching Controls.

**Supplemental Table S3** – Demographic Characteristics of Relatives of Thumb Carpometacarpal Joint Osteoarthritis Patients and Matching Controls.

**Supplemental Table S4** – Demographic Characteristics of Study Population Used for Age-Standardized Sex-Specific Incidence Rates of Thumb Carpometacarpal Joint Osteoarthritis.

**Supplemental Table S5** – Race and Ethnicity as Risk Factors for Thumb Carpometacarpal Joint Osteoarthritis.

