## Supplemental Data for "Familial Clustering and Genetic Analysis of Severe Thumb Carpometacarpal Joint Osteoarthritis in a Large Statewide Cohort"

By

Catherine M. Gavile PhD^1*^, Nikolas H. Kazmers MD, MSE^1*^, Kendra A. Novak BS^1^, Huong D. Meeks PhD^2^, Zhe Yu MS^2^, Joy L. Thomas BS^3^, Channing Hansen BS^4^, Tyler Barker PhD^1,5,6^, Michael J. Jurynec PhD^1^

^1^Department of Orthopaedics, University of Utah, Salt Lake City, UT 84108

^2^Huntsman Cancer Institute, Utah Population Database, University of Utah, Salt Lake City, UT 84112

^3^Intermountain Healthcare, Precision Genomics, St. George, UT 84790

^4^Intermountain Healthcare, Biorepository, South Salt Lake City, UT 84119

^5^Intermountain Healthcare, Precision Genomics, Murray, UT 84107

^6^Department of Nutrition and Integrative Physiology, University of Utah, Salt Lake City, UT 84112

Corresponding author: Michael J. Jurynec

Contact information:; Phone: 801-581-4424; Address: Department of Orthopaedics, University of Utah, 15 N 2030 E, Bldg 533, Salt Lake City, UT 84112

^*^C.M.G and N.H.K contributed equally to the manuscript.

**Running head:** Population and Genetic Analysis of Severe Thumb CMCJ OA

This work was funded by the Arthritis National Research Foundation, the Skaggs Foundation for Research, the Utah Genome Project, and the NIH.

**Keywords:** Carpometacarpal hand osteoarthritis, *CHSY3*, osteoarthritis gene, thumb base osteoarthritis, osteoarthritis risk factors

**Selection of Cases**

The ICD codes are provider entered based on patient visits with the possibility of the billing team reviewing and modifying for improved accuracy (https://uofuhealth.utah.edu/huntsman/utah-population-database/data/). Individuals were excluded if they were diagnosed with any of the following codes:

a.	ICD-9 codes: ICD-9 716·13, 716·14 (traumatic arthropathy, forearm and hand, respectively).

b.	ICD-10 codes: M18·2x, M18·3x (post-traumatic), M18·4 (‘other’ etiology), M18·5x (‘other’ etiology), M18·9 (unspecified etiology)

c.	Inflammatory arthritis codes: ICD9 714·0, 714·2, 714·3, ICD10 M05·xxx, M06·xx, M08·xxx.

d.	Ligamentous hyperlaxity codes: 728·4 (laxity of ligament), 718·84 (hypermobility/instability of joints, including hand), 756·83 (Ehlers-Danlos syndrome), M35·7 (hypermobility syndrome), M24·2x (ligamentous laxity NOS), Q79·6x (Ehlers-Danlos syndrome).

**Selection of Controls**

Control selection for CMCJ OA cases:

1.	Matched with cases 5:1 ratio on:

a.	Birth year

b.	Sex

c.	Have follow-up in Utah at least as long as the date of index date; controls must reside in Utah on or after the cases’ first CMCJ OA diagnosis date or CPT date, whichever is earlier.

d.	Born in/out of Utah

e.	Minimum pedigree information (e.g., if cases are singleton or have no informative relative, controls must be singleton or have no informative relative; if cases have at least one informative relative, controls must have at least one relative who is informative. Informative is defined as alive and living in Utah on/after 1/1/1996).

2.	No history of CMCJ OA (did not have CPT codes 25447 or 25445, and ICD-9 procedure codes 8269, 8174, 8175, and ICD-9/10 diagnosis codes ICD9 715·04, 715·14, M18·0, M18·1x, and M19·04)

3.	Individuals with the following codes were excluded:

a.	ICD-9 codes: ICD-9 716·13, 716·14 (traumatic arthropathy, forearm and hand, respectively).

b.	ICD-10 codes: M18·2x, M18·3x (post-traumatic), M18·4 (‘other’ etiology), M18·5x(‘other’ etiology), M18·9 (unspecified etiology).

c.	Inflammatory arthritis codes: ICD9 714·0, 714·2, 714·3, ICD10 M05·xxx, M06·xx, M08·xxx.

d.	Ligamentous hyperlaxity codes: 728·4 (laxity of ligament), 718·84 (hypermobility/instability of joints, including hand), 756·83 (Ehlers-Danlos syndrome), M35·7 (hypermobility syndrome), M24·2x (ligamentous laxity NOS), Q79·6x (Ehlers-Danlos syndrome).

**High Risk Pedigree Identification**

FSIR is a statistical method that accounts for the number of biological relatives in a pedigree, the degree of relatedness, and the age at which an individual is diagnosed^1^. Exact one-sided Poisson probabilities were calculated under the null hypothesis of no familial enrichment of CMCJ OA. Individuals were grouped into fourteen categories based on age (0-30, 31-40, 41-50, 51-60, 61-70, 71-80, and 81-120) and sex. To determine the incidence ratio, the number of years prior to and after diagnosis was calculated for all affected and unaffected individuals, and then the number of living diagnosed years was divided by the number of living undiagnosed years. To determine the pedigree incidence ratio, the UPDB was analyzed to identify the founders of pedigrees containing an affected individual, the affection status of every individual biological relative in each pedigree was determined, and incidence ratio was calculated as described above. The pedigree’s incidence ratio/whole population incidence ratio was used to determine the FSIR. High-risk pedigrees were selected if they had four or more affected individuals, and if the FSIR was ≥ 2 and significant (p < 0·05) using a chi-squared test as previously described^1^.

**Validation of Cases**

We reviewed the medical charts of 25 random cases to determine if the CMCJ OA diagnosis based on coding was correct. We were able to chart review all 25 cases. J One individual had gout, and none were diagnosed with rheumatoid arthritis, psoriasis, or psoriatic arthritis.

**Whole exome sequencing and analysis:** Whole exome sequencing (WES) and analysis was performed as previously described^2^ Briefly, WES was performed using genomic DNA isolated from saliva. We followed best practices established by the Broad Institute GATK for variant discovery (https://gatk.broadinstitute.org/hc/en-us). Analysis of variants was performed with ANNOVAR (<http://annovar.openbioinformatics.org/en/latest/>)^3^ and pVAAST (<http://www.hufflab.org/software/pvaast/>)^4^ in concert with PHEVOR2 (<http://weatherby.genetics.utah.edu/phevor2/index.html>)^5^.
