## Supplementary figures and images for "Familial Clustering and Genetic Analysis of Severe Thumb Carpometacarpal Joint Osteoarthritis in a Large Statewide Cohort"

### Supplemental Figure 1

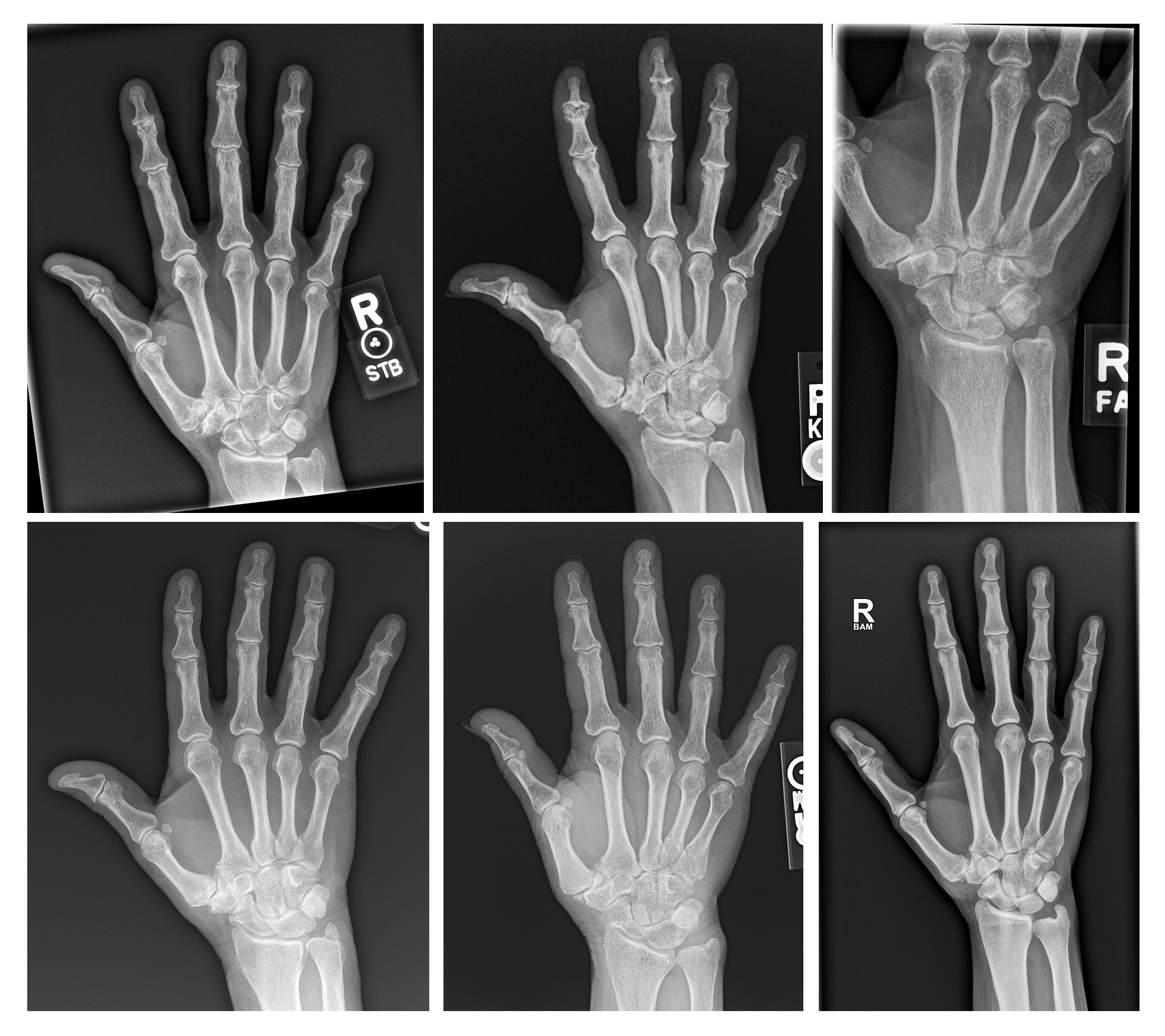
