## Supplemental Table for "Familial Clustering and Genetic Analysis of Severe Thumb Carpometacarpal Joint Osteoarthritis in a Large Statewide Cohort"

|  | Supplemental Table S1 - Identification of Risk Factors Using Diagnostic Coding |
| --- | --- |

| **Comorbidities** | **ICD-9 Diagnosis Code** | **ICD-10 Diagnosis Code** |
| --- | --- | --- |
| Alcohol related disorders | 303·x | F10·x |
| Tobacco Use | 305·1, V15·82 | F17·x, Z72·0, Z87·891 |
| Diabetes | 250·x | E08·x, E90·x, E10·x, E11·x, E13·x |
| Obesity | 278·00, 278·01, 278·03 | E66·01, E66·1, E66·2, E66·09, E66·8, E66·9 |

| Supplemental Table S2 - Demographic Characteristics of Individuals Diagnosed with Thumb Carpometacarpal Joint Osteoarthritis and Matching Controls |
| --- |

|  | **Controls** | **CMCJ OA patients** | **P-value** |
| --- | --- | --- | --- |
| N | 23,537 | 4,707 |  |
| Male (%) | 5606 (23·8) | 1124 (23·9) | 0·943 |
| Birth year (Mean (SD)) | 1950·3 (10·3) | 1950·3 (10·3) | 0·867 |
| NIH Race (%) |  |  | < 0·001 |
| - American Indian or Alaska Native | 183 ( 0·8) | 3 ( 0·1) |  |
| - Asian | 130 ( 0·6) | 6 ( 0·1) |  |
| - Native Hawaiian or Other Pacific Islander | 44 ( 0·2) | 1 ( 0·0) |  |
| - Black or African American | 93 ( 0·4) | 9 ( 0·2) |  |
| - White | 21081 (89·6) | 4091 (86·9) |  |
| - Multiple Races | 931 ( 4·0) | 82 ( 1·7) |  |
| - Unknown | 1072 ( 4·6) | 515 (10·9) |  |
| - Presumed not white, cannot be classified | 3 ( 0·0) | 0 ( 0·0) |  |
| Ethnicity (%) |  |  | < 0·001 |
| - Non-Hispanic | 17419 (74·0) | 3439 (73·1) |  |
| - Hispanic | 4177 (17·7) | 707 (15·0) |  |
| - Unknown | 1941 ( 8·2) | 561 (11·9) |  |
| Ever diagnosed with alcoholism^1^ (%) | 811 ( 3·4) | 168 ( 3·6) | 0·705 |
| Ever diagnosed with diabetes^1^ (%) | 6040 (25·7) | 1191 (25·3) | 0·619 |
| Ever diagnosed with obesity^1^ (%) | 6069 (25·8) | 1431 (30·4) | < 0·001 |
| Ever diagnosed with tobacco use^1^ (%) | 5640 (24·0) | 1369 (29·1) | < 0·001 |
| Had at least one FDR with CMC (%) | 200 ( 0·8) | 132 ( 2·8) | < 0·001 |
| Had at least one SDR with CMC (%) | 173 ( 0·7) | 43 ( 0·9) | 0·233 |
| Had at least one TDR with CMC (%) | 796 ( 3·4) | 187 ( 4·0) | 0·048 |

^1^CMC diagnoses and comorbidities diagnosis (alcohol, diabetes, tobacco, and obesity) were based on Inpatient, Ambulatory Surgery, and Intermountain Healthcare and UUHSC EDW health records between 1996 and 2020. ICD-9 and ICD-10 codes for comorbidities are listed in Supplemental Table 1.

Demographic characteristics of CMC patients and their matching controls were compared using t-tests for continuous variables and chi-square tests for categorical variables.

| Supplemental Table S3 - Demographic Characteristics of Relatives of Thumb Carpometacarpal Joint Osteoarthritis Patients and Matching Controls |
| --- |

|  | **Relatives of controls** | **Relatives of cases** | **P-values** |
| --- | --- | --- | --- |
| **First Degree Relatives (FDR)** |  |  |  |
| N | 97,353 | 18,999 |  |
| Male (%) | 48527 (49·8) | 9455 (49·8) | 0·845 |
| Birth year (Mean (SD)) | 1960·9 (22·1) | 1959·4 (22·3) | < 0·001 |
| NIH Race (%) |  |  | < 0·001 |
| - American Indian or Alaska Native | 428 (0·4) | 3 (0·0) |  |
| - Asian | 182 (0·2) | 12 (0·1) |  |
| - Native Hawaiian or Other Pacific Islander | 40 (0·0) | 3 (0·0) |  |
| - Black or African American | 84 (0·1) | 16 (0·1) |  |
| - White | 92343 (94·9) | 18365 (96·7) |  |
| - Multiple Races | 2555 (2·6) | 330 (1·7) |  |
| - Unknown | 1720 (1·8) | 270 (1·4) |  |
| - Presumed not white, cannot be classified | 1 (0·0) | 0 (0·0) |  |
| Ethnicity (%) |  |  | < 0·001 |
| - Non-Hispanic | 72963 (74·9) | 14641 (77·1) |  |
| - Hispanic | 14593 (15·0) | 2668 (14·0) |  |
| - Unknown | 9797 (10·1) | 1690 (8·9) |  |
| Diagnosed with CMC^1^ (%) | 195 (0·2) | 145 (0·8) | < 0·001 |
| **Second Degree Relatives (SDR)** |  |  |  |
| N | 230,769 | 47,027 |  |
| Male (%) | 115311 (50·0) | 23560 (50·1) | 0·609 |
| Birth year (Mean (SD)) | 1975·3 (30·6) | 1974·0 (30·8) | < 0·001 |
| NIH Race (%) |  |  | < 0·001 |
| - American Indian or Alaska Native | 808 (0·4) | 23 ( 0·0) |  |
| - Asian | 232 (0·1) | 32 ( 0·1) |  |
| - Native Hawaiian or Other Pacific Islander | 63 (0·0) | 12 ( 0·0) |  |
| - Black or African American | 177 (0·1) | 31 ( 0·1) |  |
| - White | 211212 (91·5) | 43746 (93·0) |  |
| - Multiple Races | 5154 (2·2) | 815 ( 1·7) |  |
| - Unknown | 13121 (5·7) | 2368 ( 5·0) |  |
| - Presumed not white, cannot be classified | 2 (0·0) | 0 ( 0·0) |  |
| Ethnicity (%) |  |  | < 0·001 |
| - Non-Hispanic | 164148 (71·1) | 33912 (72·1) |  |
| - Hispanic | 28931 (12·5) | 5615 (11·9) |  |
| - Unknown | 37690 (16·3) | 7500 (15·9) |  |
| Diagnosed with CMC^1^ (%) | 167 (0·1) | 44 ( 0·1) | 0·153 |
| **Third Degree Relatives (TDR)** |  |  |  |
| N | 433,398 | 95,019 |  |
| Male (%) | 219004 (50·5) | 48413 (51·0) | 0·019 |
| Birth year (Mean (SD)) | 1973·6 (31·4) | 1973·1 (31·6) | < 0·001 |
| NIH Race (%) |  |  | < 0·001 |
| - American Indian or Alaska Native | 1170 (0·3) | 54 ( 0·1) |  |
| - Asian | 256 (0·1) | 38 ( 0·0) |  |
| - Native Hawaiian or Other Pacific Islander | 49 (0·0) | 14 ( 0·0) |  |
| - Black or African American | 287 (0·1) | 58 ( 0·1) |  |
| - White | 392647 (90·6) | 86836 (91·4) |  |
| - Multiple Races | 7721 (1·8) | 1430 ( 1·5) |  |
| - Unknown | 31261 (7·2) | 6589 ( 6·9) |  |
| - Presumed not white, cannot be classified | 6 (0·0) | 0 ( 0·0) |  |
| Ethnicity (%) |  |  | < 0·001 |
| - Non-Hispanic | 317216 (73·2) | 70190 (73·9) |  |
| - Hispanic | 49778 (11·5) | 10342 (10·9) |  |
| - Unknown | 66404 (15·3) | 14487 (15·2) |  |
| Diagnosed with CMC^1^ (%) | 775 (0·2) | 207 ( 0·2) | 0·013 |

^1^Diagnosed with CMC based on Inpatient, Ambulatory Surgery, and Intermountain Healthcare and UUHSC EDW health records between 1996 and 2020.

Demographic characteristics of relatives of CMC patients and their matching controls were compared using t-tests for continuous variables and chi-square tests for categorical variables.

| Supplemental Table S4 - Demographic Characteristics of Study Population Used for Age-Standardized Sex-Specific Incidence Rates of Thumb Carpometacarpal Joint Osteoarthritis |
| --- |

|  | **No CMCJ OA diagnosis** | **Diagnosed with CMCJ OA** | **P-values** |
| --- | --- | --- | --- |
| N | 6,083,729 | 4,540 |  |
| Male (%) | 3,121,384 (51·3%) | 1,077 (23·7%) | <0·001 |
| Birth year (Mean (SD)) | 1980·2 ± 24·6 | 1950·3 ± 10·3 | <0·001 |
| Year of 1^st^ CMC diagnosis reported | - | 2012·8 ± 5·6 | - |
| - 1996 | - | 10 (0·2) |  |
| - 1997 | - | 22 (0·5) |  |
| - 1998 | - | 52 (1·1) |  |
| - 1999 | - | 31 (0·7) |  |
| - 2000 | - | 60 (1·3) |  |
| - 2001 | - | 72 (1·6) |  |
| - 2002 | - | 91 (2·0) |  |
| - 2003 | - | 93 (2·0) |  |
| - 2004 | - | 95 (2·1) |  |
| - 2005 | - | 139 (3·1) |  |
| - 2006 | - | 123 (2·7) |  |
| - 2007 | - | 106 (2·3) |  |
| - 2008 | - | 130 (2·9) |  |
| - 2009 | - | 146 (3·2) |  |
| - 2010 | - | 145 (3·2) |  |
| - 2011 | - | 157 (3·5) |  |
| - 2012 | - | 179 (3·9) |  |
| - 2013 | - | 230 (5·1) |  |
| - 2014 | - | 235 (5·2) |  |
| - 2015 | - | 321 (7·1) |  |
| - 2016 | - | 538 (11·9) |  |
| - 2017 | - | 664 (14·6) |  |
| - 2018 | - | 669 (14·7) |  |
| - 2019 | - | 141 (3·1) |  |
| - 2020 | - | 83 (1·8) |  |
| - 2021 | - | 8 (0·2) |  |

Demographic characteristics of CMC patients and the population without any CMC diagnosis in Utah from 1996-2020 were compared using t-tests for continuous variables and chi-square tests for categorical variables.

| Supplemental Table S5 - Race and Ethnicity as Risk Factors for Thumb Carpometacarpal Joint Osteoarthritis |
| --- |

**Both sexes – adjusted for diabetes**

|  | P-value | Relative Risk | 95% CI Lower Limit | 95% CI Upper Limit |
| --- | --- | --- | --- | --- |
| Race (Non-white vs. White) | 0·000 | 0·41 | 0·33 | 0·51 |
| Race (Unknown vs. White) | 0·000 | 4·80 | 3·95 | 5·83 |
| Ethnicity (Hispanic vs. Non-Hispanic) | 0·000 | 0·85 | 0·78 | 0·93 |
| Ethnicity (Unknown vs. Non-Hispanic) | 0·000 | 0·69 | 0·59 | 0·82 |

**Males – adjusted for diabetes**

|  | P-value | Relative Risk | 95% CI Lower Limit | 95% CI Upper Limit |
| --- | --- | --- | --- | --- |
| Race (Non-white vs. White) | 0·002 | 0·52 | 0·34 | 0·79 |
| Race (Unknown vs. White) | 0·000 | 5·32 | 3·48 | 8·13 |
| Ethnicity (Hispanic vs. Non-Hispanic) | 0·846 | 0·98 | 0·81 | 1·19 |
| Ethnicity (Unknown vs. Non-Hispanic) | 0·049 | 0·70 | 0·48 | 1·00 |

**Females – adjusted for diabetes**

|  | P-value | Relative Risk | 95% CI Lower Limit | 95% CI Upper Limit |
| --- | --- | --- | --- | --- |
| Race (Non-white vs. White) | 0·000 | 0·39 | 0·31 | 0·49 |
| Race (Unknown vs. White) | 0·000 | 4·65 | 3·74 | 5·79 |
| Ethnicity (Hispanic vs. Non-Hispanic) | 0·000 | 0·82 | 0·74 | 0·91 |
| Ethnicity (Unknown vs. Non-Hispanic) | 0·000 | 0·69 | 0·57 | 0·83 |

**Both sexes – not adjusted for diabetes**

|  | P-value | Relative Risk | 95% CI Lower Limit | 95% CI Upper Limit |
| --- | --- | --- | --- | --- |
| Race (Non-white vs. White) | 0·000 | 0·41 | 0·33 | 0·50 |
| Race (Unknown vs. White) | 0·000 | 4·80 | 3·96 | 5·83 |
| Ethnicity (Hispanic vs. Non-Hispanic) | 0·000 | 0·85 | 0·77 | 0·93 |
| Ethnicity (Unknown vs. Non-Hispanic) | 0·000 | 0·70 | 0·59 | 0·82 |

**Males – not adjusted for diabetes**

|  | P-value | Relative Risk | 95% CI Lower Limit | 95% CI Upper Limit |
| --- | --- | --- | --- | --- |
| Race (Non-white vs. White) | 0·002 | 0·52 | 0·34 | 0·79 |
| Race (Unknown vs. White) | 0·000 | 5·31 | 3·47 | 8·10 |
| Ethnicity (Hispanic vs. Non-Hispanic) | 0·865 | 0·98 | 0·81 | 1·19 |
| Ethnicity (Unknown vs. Non-Hispanic) | 0·044 | 0·69 | 0·48 | 0·99 |

**Females – not adjusted for diabetes**

|  | P-value | Relative Risk | 95% CI Lower Limit | 95% CI Upper Limit |
| --- | --- | --- | --- | --- |
| Race (Non-white vs. White) | 0·000 | 0·38 | 0·30 | 0·49 |
| Race (Unknown vs. White) | 0·000 | 4·66 | 3·74 | 5·81 |
| Ethnicity (Hispanic vs. Non-Hispanic) | 0·000 | 0·81 | 0·73 | 0·90 |
| Ethnicity (Unknown vs. Non-Hispanic) | 0·000 | 0·69 | 0·58 | 0·84 |

CMC and comorbidities diagnosis were based on Inpatient, Ambulatory Surgery, and Intermountain Healthcare and UUHSC EDW health records between 1996 and 2020.
